# Gastric Alimetry^®^ impacts the management pathway of chronic gastroduodenal disorders

**DOI:** 10.1101/2023.02.06.23285567

**Authors:** Charlotte Daker, Chris Varghese, William Xu, Chris Cederwall

## Abstract

**Background:** Gastric Alimetry is a new diagnostic tool using non-invasive gastric electrical mapping and symptom logging to identify patient subgroups. This study aimed to propose an initial framework for Gastric Alimetry implementation in the routine management of gastroduodenal disorders, and assess its impact on diagnosis and management.

**Methods:** Gastric Alimetry using standard tests (30-min fasted, ~480kCal meal, followed by 4-hr postprandial recording with concurrent symptom logging) was applied to patients presenting with gastroduodenal symptoms.

**Results:** Overall, 50 patients were evaluated with Gastric Alimetry. The test aided management decisions in 78% of patients (39/50) and aided a change in diagnosis in 40% (20/50), predominantly from a motility disorder to disorders of gut-brain interaction (DGBI). Changes in invasive nutritional support occurred in 18% (9/50).

**Conclusion:** Gastric Alimetry impacted care in most patients in this first series. Further work to inform clinical utility is now a priority.

## Introduction

Chronic gastroduodenal symptoms pose a diagnostic and therapeutic challenge. These symptoms are commonly seen in disorders of gut-brain interaction (DGBI), including functional dyspepsia (FD) and chronic nausea and vomiting syndrome (CNVS), as well as in gastroparesis with delayed gastric emptying.(1,2) However, recent evidence shows these disorders are clinically interchangeable because of overlapping symptom profiles and tests.(3) More specific diagnostic pathways are therefore required to guide targeted care.(4)

Gastric Alimetry® (Alimetry, New Zealand) is a new diagnostic device, combining non-nvasive gastric electrical mapping with simultaneous symptom logging in a validated App.(5,6). The gastric mapping data is capable of identifying patient subgroups with neuromuscular dysfunction, while the symptom data can help determine symptom origins.(5,6)

Gastric Alimetry is now being introduced into practice, challenging existing models of care. As such, the current study had two aims. First, to propose an initial framework for the implementation of Gastric Alimetry in the routine management of gastroduodenal disorders, and second to assess its impact on diagnosis and management in a consecutive series of 50 cases.

## Methods

The new management framework is presented in **Figure 1A**, with justification below. The framework was formulated to integrate Gastric Alimetry, gastric emptying test (GET), and existing clinical guidelines. The model was applied pragmatically with reference to individual patient histories, clinical work-up, and comorbidities, while also accepting that gastroduodenal disorders and phenotypes may overlap.(2)

**Figure 1.**
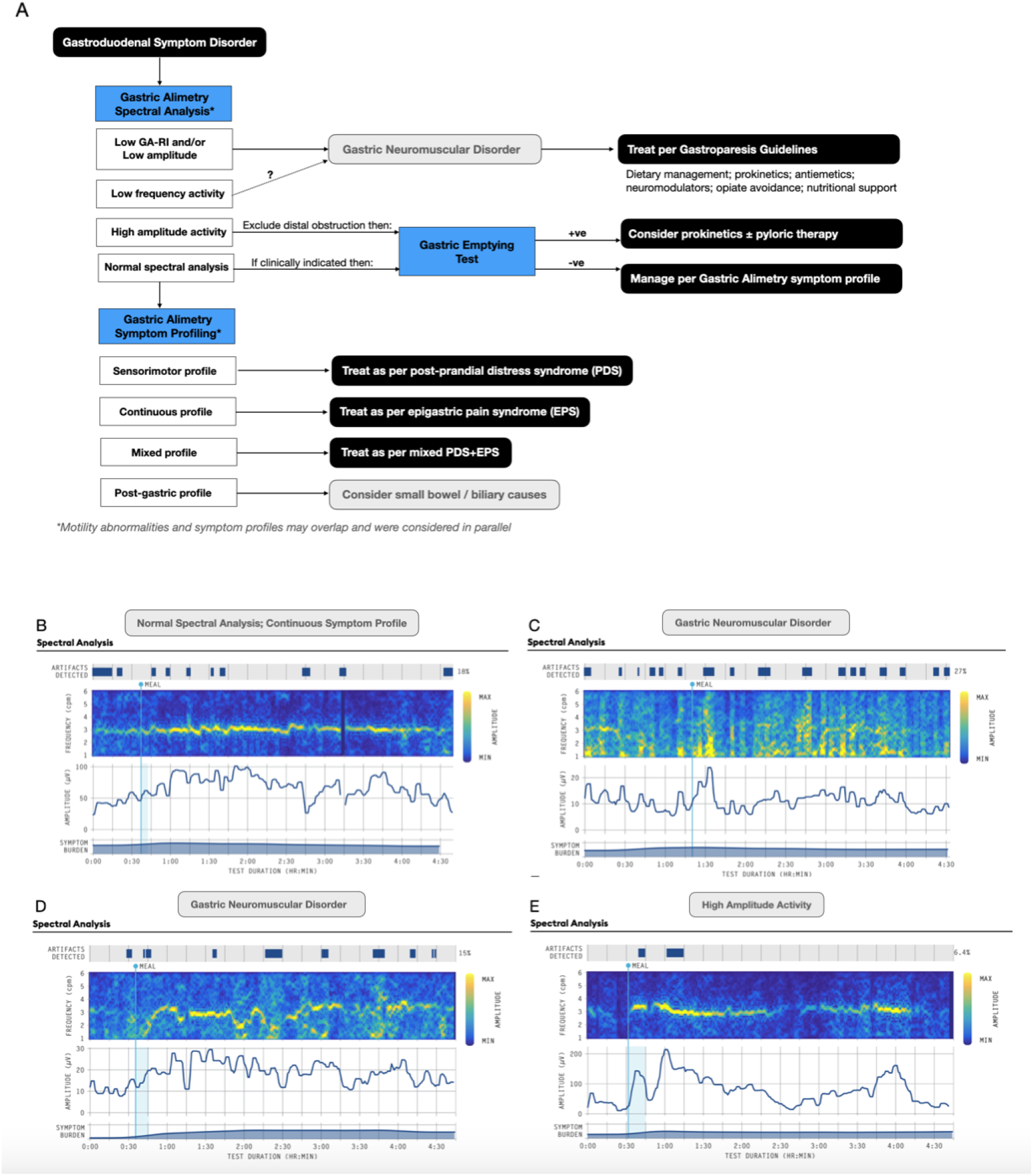
**A**) Flow diagram developed for the application of Gastric Alimetry in the management of gastroduodenal disorders. Patients may have more than one phenotype. **B-D**) Examples of Gastric Alimetry spectral data, amplitude plots, and symptom burden graphs from the study cohort. **B)** *Normal spectral analysis; continuous symptom profile*: Principal Gastric Frequency (PGF) 3.01 cpm (range 2.65-3.35); Gastric Alimetry Rhythm Index 0.88 (GA-RI; range ≥0.25); BMI-adjusted amplitude 61.3±17.8 μV (range 22-70 μV). **C)** *Neuromuscular profile* (GA-RI 0.12; 21.6 μV); **D)** *Neuromuscular profile with a sensorimotor symptom profile* (GA-RI 0.12; 21.6 μV); **E)** *High amplitude activity (*average 72.1 μV).

### • Gastric Alimetry Spectral Analysis

provides the following major phenotypes:

i. *Gastric neuromuscular disorder:* Gastric Alimetry Rhythm Index (GA-RI) <0.25, and/or low amplitudes <22 μV.(7) Managed as per gastroparesis guidelines,(1) regardless of GET status.
ii. *High sustained BMI-adjusted amplitude*: >70 μV.(7) Consider distal obstruction, and evaluate GET with consideration of pyloric therapies.(8)
iii. *Normal spectral activity*: Request GET if indicated.(1) If delayed, prokinetics initiated and pyloric therapies considered;(1) if normal, managed per symptom profile below.

### • Gastric Alimetry Symptom Profiling

provides the following major phenotypes:

i. *Sensorimotor:* symptoms correlate with gastric amplitude, suggesting hypersensitivity / accommodation disorders. Consider therapy per post-prandial distress syndrome.(2)
ii. *Continuous profile*: symptoms constant / do not correlate with gastric amplitude; consider therapy per epigastric pain syndrome.(2)
iii. *Mixed / other:* combination or absence of above profiles. Managed per clinical impression.
iv. *Post-gastric*: symptoms trending upward late in post-prandial period; investigate more distal disorders.

The clinical impact of Gastric Alimetry was evaluated based on a study addressing the impact of antroduodenal manometry.(9) Charts were reviewed by an independent assessor, with a positive management impact defined as an outcome that established a new diagnosis or altered therapy (medication, endoscopic intervention, feeding).(9) Ethical approval was obtained.

## Results

A total of 50 patients were evaluated (Wellington / North Shore Hospitals, New Zealand). All patients underwent standard Gastric Alimetry tests: 30-min fasted, ~480kCal meal, followed by 4-hr postprandial recording with concurrent symptom logging.(5,6) **Figure 1B**. shows representative examples of phenotypes encountered in the cohort.

Of the 50 patients, 45 were female (90%); median age 30 years (IQR = 22-46). Predominant presenting symptoms were nausea/vomiting (72%), upper abdominal pain (8%), other functional gastrointestinal symptoms (e.g. bloating, fullness, heartburn) (20%). The majority had a previous GET (70%). Primary diagnoses prior to Gastric Alimetry evaluation were gastroparesis / other gut motility disorder (23/50; 46%), or DGBI (CNVS / FD) in 27/50 (54%).

The diagnostic outcomes of the Gastric Alimetry tests are summarized in **Figure 2A**, with impact on diagnosis and management summarized in **Figures 2B-C**. Overall, the test aided a management decision in 39/50 patients (78%). A significant change in diagnosis occurred 40% (20/50), typically being a switch from a primary motility disorder to a primary DGBI / sensory disorder, or vice-versa, as based on the spectral analysis (**Figure 2A**). Therapeutic changes favored prokinetics in 28%, neuromodulators in 36%, with two patients receiving pyloric intervention, and management focused predominantly on other medications in 10% (**Figure 2B**).

**Figure 2.**
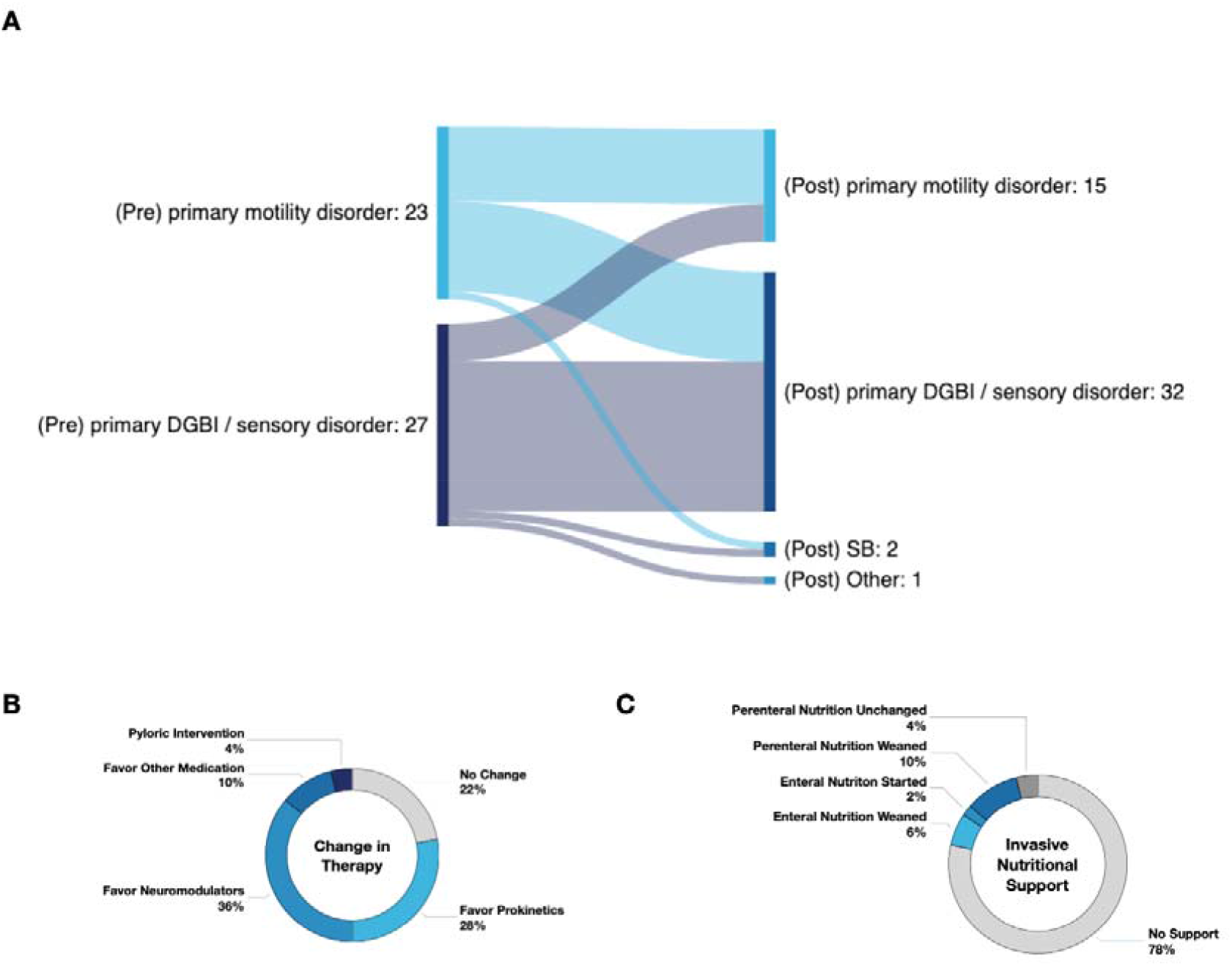
**A**) Sankey diagram showing the changes in diagnoses that occurred between a primary motility / neuromuscular disorder and DGBI/sensory disorders (pre Gastric Alimetry = left column) vs *post Gastric Alimetry* diagnoses (right column). **B)** Significant changes in management that were guided by Gastric Alimetry test data, as per the workflow in Figure 1A. **C)** Significant changes in invasive (parenteral or enteral nutrition) that were achieved as a result of the changes instituted in Figure 2A,B in tandem with integrated clinical care.

Changes in invasive nutritional support occurred in 9/50 cases (18%) (**Figure 2C**). Of 7 patients on parenteral nutrition prior to evaluation, 5 were successfully weaned, together with 3 patients on enteral feeding, after management changes facilitated tolerance of diet. Conversely, nasojejunal feeding was initiated in one patient with a neuromuscular disorder who failed to respond to medical management.

## Discussion

These data present the first real-world report of the introduction of Gastric Alimetry into a clinical care pathway. The results showed that a high majority (78%) of patients achieved a positive impact from the new test, with the most consequential changes being a switch from a motility disorder to DGBI / sensory diagnosis (40% of patients). These changes were justified because GET is now established to be insensitive and non-specific for neuromuscular abnormalities, whereas Gastric Alimetry is targeted toward neuromuscular profiling.(3,5,7) It should also be noted that the test was only an aid to decision making, and clinicians also considered other patient inputs at test follow-up that likely contributed to the high rate of changes in clinical care.

These results offer an alternative paradigm in managing gastroduodenal symptoms. In particular, the framework reduces reliance on symptom criteria and GET, which present overlapping profiles,(3) and instead proposes phenotyping patients based on specific mechanisms when possible.(5–7) The phenotypes applied here cover several known gastroduodenal pathologies, including neuromuscular disorders (including interstitial cell of Cajal pathologies), visceral hypersensitivity, DGBI, and gastric outlet resistance. While these disorders are well established, they have previously been challenging to separate and apply at the individual patient level. Mechanistic phenotyping then facilitated personalized therapy, including earlier and more aggressive approaches to second or third-line neuromodulator or prokinetic agents in selected patients, while reducing trial-and-error.

These changes in care pathways may have substantial implications for outcomes, costs of care, and quality of life in gastroduodenal disorders. This was highlighted by several patients being weaned from parenteral nutrition in this cohort. However, it is also emphasized that this study only reports initial diagnostic and management changes, and prospective studies showing additional patient outcome data are now a priority. In addition, as Gastric Alimetry is a new test, we anticipate that the new model will be refined and improved as use of the device expands.

## Data Availability

All data produced in the present study are available upon reasonable request to the authors

## Notes

**Conflicts of interest:** The authors have no conflicts of interest to declare.

**Funding statement:** This project was supported by the Health Research Council of New Zealand.

### Competing Interest Statement

The authors have declared no competing interest.

### Funding Statement

The Health Research Council of New Zealand funded this work

### Author Declarations

Auckland Health Research Ethics Committee of University of AUckland gave ethical approval for this work

